# Inhibition of BIRC5 and MCL1 as a potential treatment strategy to overcome drug resistance in Mantle Cell Lymphoma

**DOI:** 10.1101/2025.04.10.25322797

**Authors:** Jeremiah Pfitzer, Sayak Chakravarti, Suman Mazumder, Fengzhi Li, Ujjal Kumar Mukherjee, Amit Kumar Mitra

## Abstract

Mantle cell lymphoma (MCL) is a difficult-to-cure, highly heterogeneous, and aggressive form of non-Hodgkin lymphoma with a high recurrence rate and poor long-term prognosis. Using a novel optimization-regularization-based computational prediction algorithm called “secDrug,” we have identified therapeutic targets for treating resistant cancers. When applied to B-cell malignancies, the secDrug algorithm predicted the pro-survival proteins BIRC5 and MCL1 as top targets. Here, using a panel of MCL cell lines representing PI/BTKi sensitive, innate resistance, and clonally derived acquired resistance, we demonstrated that small molecule-based inhibition of BIRC5 and MCL1 is effective in killing MCL cells as single agents and in combination with Bortezomib or Ibrutinib. Further, using bulk and single-cell transcriptomics, gene knockdown studies, and cell-based assays, we identified genes and molecular networks associated with their mechanism of action and synergy. *In silico* analysis using patient datasets underlined the clinical potential of these candidates in curbing MCL progression and drug resistance.

## INTRODUCTION

Mantle cell lymphoma (MCL) is an aggressive lymphoid neoplasm that develops from malignant B-lymphocytes in the outer edge or mantle zone of a lymph node^1^. MCL is a subtype of B-cell non-Hodgkin lymphoma characterized by rapid clinical progression and poor response rate to conventional chemotherapeutic drugs, with recurrent relapse resulting in a short estimated 5-year overall survival (OS) of 2-5 years^2,3^. Despite promising initial responses to first-line MCL therapies, almost all patients develop resistance over time^4,5^. The Bruton’s tyrosine kinase inhibitor (BTKi) Ibrutinib (IBR) and the proteasome inhibitor (PI) Bortezomib (BTZ) are FDA-approved therapies for MCL with demonstrated high initial response rate in clinical trials. ^6,7^. However, the majority of these patients either have an innate resistance to these therapies or eventually acquire resistance over the course of treatment, thus progressing into more aggressive incurable refractory or relapsed (R/R) disease states, with a median progression-free survival (PFS) of <15 months^3,8,9^. In this context, the identification of novel drugs that function either alone or in combination to curb the oncogenic progression is of high clinical significance. For this purpose, we have designed a novel pharmacogenomics data-driven optimization-regularization-based computational prediction algorithm to predict molecular targets for the development of small molecule inhibitors as novel secondary drug candidates (’secDrugs’) for the management of treatment-resistant cancers^10^. Previously, we demonstrated that the top-predicted secDrug candidates show significant cytotoxicity against *in vitro* and *ex vivo* (patient-derived tumor cells) models of advanced-stage cancers, including relapsed/resistant multiple myeloma (R/R MM) and lethal variants of Prostate Cancer, either alone or in combination with primary drugs^10,11^. When applied to drug-resistant B-cell malignancies, our algorithm predicted BIRC5 and the pro-apoptotic protein MCL1 as promising therapeutic secDrug targets in R/R MCL. Several lines of evidence implicate dysfunctional Inhibitors of Apoptosis Proteins (IAPs) like BIRC5 in R/R MCL, thus signifying the need for investigation into IAP-targeted therapy. MCL1, a member of the BCL-2 family, is associated with poor outcomes in the treatment of B-cell cancers^12,13^. Interestingly, BIRC5 and MCL1 have both been reported to be over-expressed in MCL and strongly correlated with oncogenic progression and survivability of MCL patients^14–17^. We hypothesize that a combination of the inhibition of these targets using MCL1 (BIRC5 inhibitor) and S63845 (MCL1 inhibitor) with BTKi and PI will be useful in curbing oncogenic progressions of MCL and abrogating drug resistance through simultaneous inhibition of multiple pathways.

## MATERIALS AND METHODS

### Drugs and Reagents

Drugs, reagents, antibodies, and kits are listed in **Table S1**. All the drugs were initially dissolved in dimethyl sulfoxide (DMSO, Sigma-Aldrich; St. Louis, MO, US) and stored at -20 °C.

### Human cell lines

MCL cell lines (MINO, JEKO1, Z-138) and the human stromal cell line HS-5 were obtained from ATCC (Manassas, VA). All the cell lines were cultured in media recommended by the supplier and were maintained in an incubator at 37°C with 5% CO_2_. The cell lines were authenticated at the source and tested at regular intervals for mycoplasma negativity.

### Establishment of acquired resistant MCL cell line

PI-resistant MINO-VR was created from the clonally related parental PI-sensitive MINO-P MCL cell line by dose escalation (**Figure S2**). Briefly, the MINO-P cell line was subjected to pulses of once-weekly Bortezomib treatment. Bortezomib concentrations were doubled after every three weeks of treatment. The process of dose escalation continued for six months. Cultures were removed from bortezomib for 14 days or 6 months before analysis and cultured in a manner consistent with the parental lines.

### *In vitro* chemosensitivity assays and drug synergy analysis

MCL cell lines were treated with serial dilutions of the ‘secDrugs’ - YM155 and S63845, primary drugs - PI (represented by BTZ) and BTKi (represented by Ibrutinib), and FL118 as single agents or in combination for 48hr, and cytotoxicity assays were performed using CellTiter-Glo Luminescent cell viability assay (Promega Corporation, Madison, WI). Luminescence was measured by BioTek Synergy Neo2 Microplate Reader (Winooski, VT, US). Half-maximal inhibitory concentration (IC_50_) values were determined by calculating the nonlinear regression using a sigmoidal dose-response equation (variable slope).

For combination therapy, cell lines were treated with an indicated concentration of PI and BTKi and two doses of either YM155 (5 and 10 nM) or S63845 (250 nM and 2.5 µM) for 48hr. The combination Index (CI) of each treatment was calculated in Calcusyn Software (Biosoft) using Chou-Talalay’s Median Effect method^18,19^. CI<1 depicts synergism, CI=1 refers to additive effect, and CI>1 depicts antagonism.

### Apoptosis assays

MCL cells were treated with single-agent primary drugs and secDrugs as well as combination of drugs (primary drug + secDrug), harvested and washed, followed by staining in the dark with Annexin V-FITC and Propidium Iodide according to manufacturer’s protocol (BD; Franklin Lakes, NJ). Data was acquired by BD LSR II flow cytometry (BD; Franklin Lakes, NJ) and analyzed in FlowJo™ Software (Ashland, OR).

Caspase-3/7 activity assay was performed on the MCL cell lines using Caspase-Glo 3/7 luminescent assay kit according to manufacturer’s instructions (Promega Corporation, Madison, WI). Luminescence was measured using BioTek Synergy Neo2 microplate reader. Caspase activity was normalized to untreated controls and plotted using GraphPad Prism (LaJolla, CA).

### Aldeflour Activity Assay

Aldehyde dehydrogenase (ALDH) activity was assessed using the ALDEFLUOR kit according to the manufacturer’s instructions (Stem Cell Technologies; Vancouver, Canada). Briefly, primary drug and secDrug single-agent treated MCL cells were harvested and resuspended in ALDEFLUOR assay buffer containing the ALDH substrate, BODIPY-amino acetaldehyde. Negative control samples were treated with diethylamino benzaldehyde (DEAB), an inhibitor of ALDH1 enzymatic activity. Then, the MCL cells were suspended in ALDEFLUOR assay buffer, and the brightly fluorescent ALDH^+^ cells were detected by BD LSR II flow cytometer (BD; Franklin Lakes, NJ) and analyzed using FlowJo™ Software (Ashland, OR).

### Mitochondrial transmembrane potential measurement (ΔΨ m)

Mitochondrial membrane potential was measured using the JC-1 Mitochondrial Membrane Potential Assay Kit by following the manufacturer’s protocol (Abcam, Cambridge, UK). Briefly, the cells were plated in a 96-well black/clear-bottom plate and treated with 0.1% DMSO or the drugs (primary drugs, secDrug-single-agent, and combination). Following incubation, the cells were stained with JC-1 dye, and fluorescence was recorded in a Synergy Neo 2 Microplate Reader (BioTek; Winooski, VT, US).

### Depletion of target genes by siRNA

siRNA knockdown was performed to evaluate *in vitro* cytotoxicity in target gene-depleted cells following BTZ and IBR treatment. Briefly, 27-mer TriFECTa dicer-substrate RNA duplex (DsiRNAs), positive control, and negative control (DS NC-1) DsiRNAs were purchased from IDT. These DsiRNAs were reverse-transfected in MCL cell lines with Lipofectamine RNAiMAX Transfection Reagent (Invitrogen™) in Opti-MEM I Reduced Serum Medium (Gibco) and incubated for 10 minutes at room temperature. Then, 20 µL/96-well, 100 µL/24-well, and 500 µL/6-well were added to each well of the experimental plates, and the cells were passaged. For cell viability assays, cells were resuspended in antibiotic-free media, and 10,000 cells/96-well and 250,000 cells/6-well were added to the plates. Bortezomib (final concentrations: 1 nM, 2 nM, 4 nM) or Ibrutinib (final concentrations: 1 µM, 5 µM, 10 µM, and 20 µM) were added to the wells, and cells were incubated at 37°C for 24h. *In vitro* cytotoxicity was assessed with CellTiter-Glo Luminescent Cell Viability Assay at 24h and 48h (Promega) as described above.

### Quantitative estimation of target gene expression using quantitative real-time PCR (qPCR)

Downregulation of gene expression following siRNA knockdown was assessed via quantitative reverse-transcriptase PCR (RT-qPCR). Total RNA was extracted from MCL cells using the QIAshredder and RNeasy RNA isolation kit. RNA concentration was determined using the Nanodrop-8000 spectrophotometer, and cDNA was synthesized using the QuantiTect reverse transcription kit (Qiagen). qPCR was run using gene-specific TaqMan Assay probes on a CFX96 Touch Real-Time PCR Detection System (BioRad) following the manufacturer’s protocol. Reactions were run in triplicates, and Cq values were estimated using CFZ Maestro software (BioRad). Relative gene expression was calculated using the 2^-ΔΔCt^ method following normalization with Beta-Actin expression (housekeeping gene). The fold change in target gene expression was then calculated and compared with the negative control.

### Single-cell gene expression analysis (scRNAseq)

Untreated MinoP and Mino-VR were subjected to automated single-cell capture and cDNA synthesis using the 10X Genomics (Pleasanton, CA, US) Chromium platform. Single-cell RNA sequencing (scRNA-seq-based gene expression analysis) was performed on the Illumina HiSeq 2500 Next-generation sequencing platform (Paired-end, 2*125bp, 100 cycles, v3 chemistry) at ∼5 million reads per sample.

scRNAseq datasets were obtained as matrices in the Hierarchical Data Format (HDF5 or H5). We used a combination of Seurat, CellRanger, and Partek Flow software packages to pre-process scRNA-seq data and perform single-cell gene expression analysis for biomarker-based identification of PI/BTKi-resistant single-cell subpopulations, subclones expressing cancer stem-like signatures, as well as secDrug treatment-induced erosion of these subclones. Highly variable genes will be selected for clustering analysis based on a graph-based clustering approach. t-distributed stochastic neighbor embedding (t-SNE) and UMAP (Uniform Manifold Approximation and Projection) plots were generated to visualize the cell subpopulation architecture based on markers of interest. Relative marker intensities and cluster abundances per sample were visualized by a heatmap.

### Whole-transcriptome gene expression analysis (RNAseq)

MCL cells were plated and incubated overnight at 37°C in a 6-well plate, followed by treatment with YM155 and S63845 as single agents and in combination with PI and BTKi. After 24hr, cells were harvested, and high-quality RNA was extracted using QIAshredder and RNeasy kit (Qiagen; Hilden, Germany) and stored at -80°C. RNA concentration was measured using a Nanodrop-8000 spectrophotometer (Thermo-Fisher Scientific; Waltham, MA, US), and RNA integrity was assessed using an Agilent 2100 Bioanalyzer (Agilent Technologies; Santa Clara, CA, US). An RNA integrity number (RIN) threshold >8 was applied, and RNA-seq libraries were constructed using Illumina TruSeq RNA Sample Preparation kit v2 (Illumina; San Diego, CA). The libraries were then size-selected, and RNA sequencing was performed on Illumina’s NovaSeq platform using a 150 bp paired-end protocol with a depth of > 20 million reads per sample.

### RNAseq data analysis

RNAseq data obtained as fastq files was pre-processed and mapped to the most recent human genome build Hg38 using STAR aligner. Gene counts were generated using the Ensembl annotation database (Ensembl Transcripts release 113), and annotated genes with mean counts <10 were removed. The filtered data was normalized, and differential gene expression analysis was performed between two groups of RNAseq datasets (e.g., treated vs. untreated) using a combination of command-line-based analysis pipeline and Partek Flow software package (Partek, Inc.; St. Louis, MO, US). We used gene-specific analysis (GSA) to analyze differential gene expression between groups that applies limma, an empirical Bayesian method that increases statistical power, to detect the differentially expressed genes (DEG). Multiple testing was performed using the Benjamini–Hochberg method. Mean fold-change > |1| and p < 0.05 were considered as the threshold for reporting significant differential gene expression. Heatmaps were generated using unsupervised hierarchical clustering (HC) analysis based on the top DEGs.

### Pathway analysis

The significant differentially expressed genes (DEGs) were used to perform pathway analysis using the Ingenuity Pathway Analysis (IPA) software (Qiagen; Hilden, Germany) to identify i) the most significantly affected molecular pathways associated with the mechanism of action (MoA) and mechanism of response (MoR), ii) upstream regulator molecules, iii) downstream effects and biological processes, and iv) causal networks, predicted to be activated or inhibited in response to secDrug single agent and combination treatments.

### Western Blotting

MCL cells were treated with respective drugs at indicated concentrations and were harvested, washed, and lysed using cell lysis buffer containing 50□mM Tris-HCl, pH 7.5, 150□mM NaCl, 1% NP40, 5□mM EDTA, 1□mM DTT, phosphatase, and protease inhibitors cocktail (Sigma) and incubated on ice for 10 minutes. The samples were centrifuged at 13,500□rpm at 4°C for 30□minutes, and the supernatant was collected and quantified using Pierce™ BCA Protein Assay Kit (Thermo-Fisher Scientific; Waltham, MA). Samples were prepared in reducing sample buffer and loaded on 4-12% polyacrylamide gels (Mini-PROTEAN TGX, Bio-Rad 4-15%) and then transferred onto PVDF membranes (Millipore; Billerica, MA). Membranes were blocked with 5% BSA for 1hr at room temperature, followed by overnight incubation with primary antibodies (1:1000 dilution) at 4^°^C. Next day, the blots were washed three times with TBS-0.1% Tween 20 buffer (TBS-T) and incubated with appropriate secondary antibody (1:10,000 dilution) at room temperature for 1hr. After 4x wash with TBS-T buffer, the blots were developed by adding chemiluminescent HRP substrate (Bio-Rad Laboratories; Hercules, CA), and the exposed image was captured using a ChemiDoc™ MP Imaging System (Bio-Rad). Densitometry analysis was performed using Image Lab software (BioRad).

### Patient datasets

Gene expression data on patients with lymphoid B-cell malignancies was extracted from The Cancer Genome Atlas (TCGA) database Genomic Data Commons (GDCs) server (https://www.cancergenome.nih.gov). The web-based tool Gene Expression Profiling Interactive Analysis (GEPIA) was then used to compare the expression of top genes identified through differential gene expression analysis with patient survival (overall survival (OS)/disease-free survival (DFS))^20,21^. Further, the GSE141335 dataset that contains expression data on primary samples from mantle cell lymphoma patients (n=60) was used to screen the top DE genes for association with *ex vivo* response to Ibrutinib treatment^22^.

### Statistical analysis

All statistical analyses were performed using R software environment for statistical computing and graphics, as well as GraphPad Prism. All tests were two-sided, and p<0.05 was considered statistically significant. Multiple testing was performed using the Benjamini–Hochberg method.

## RESULTS

### Identification of secondary drugs using the secDrug algorithm

The design and development of our pre-clinical drug development pipeline called “secDrug” is described earlier^10^. Briefly, the secDrug algorithm incorporates a novel, data-driven, modified greedy algorithm/minimal set-cover/computational optimization method followed by regularization to seek gene/pathway targets for secondary drug candidates (’secDrugs) that could kill the maximum number of drug-resistant B-Cell cancer lines in a sequential manner ordered by the number of cell lines killed ^10^ The top among the predicted secondary drug targets were BIRC5 and MCL1. Therefore, we selected YM155 (a survivin/BIRC5 inhibitor) and S63845 (a BH3 mimetic and an inhibitor of MCL1) for our proof-of-concept experiments in MCL.

### Single Cell transcriptomics (scRNAseq)-based secDrug screening

First, we used scRNAseq as a novel biomarker-based drug screen to identify single-cell sub-clones (represented by t-SNE clusters) in the untreated drug-sensitive and resistant MCL cell lines representing sensitive, relapse, and/or refractory MCL that harbor secDrug target genes. **Figure 1A-C** shows a representative t-SNE plot for the MINO sensitive/resistant pair (n=3841 cells) where our scRNAseq data demonstrated that the majority of the single-cell clusters have high expression of BIRC5, the S63845 target gene MCL1, and a common upstream gene (*DDX5)* indicating that the secDrugs YM155, a known survivin/BIRC5 inhibitor, and S63845 may be effective against these subpopulation clusters.

**Figure 1.**
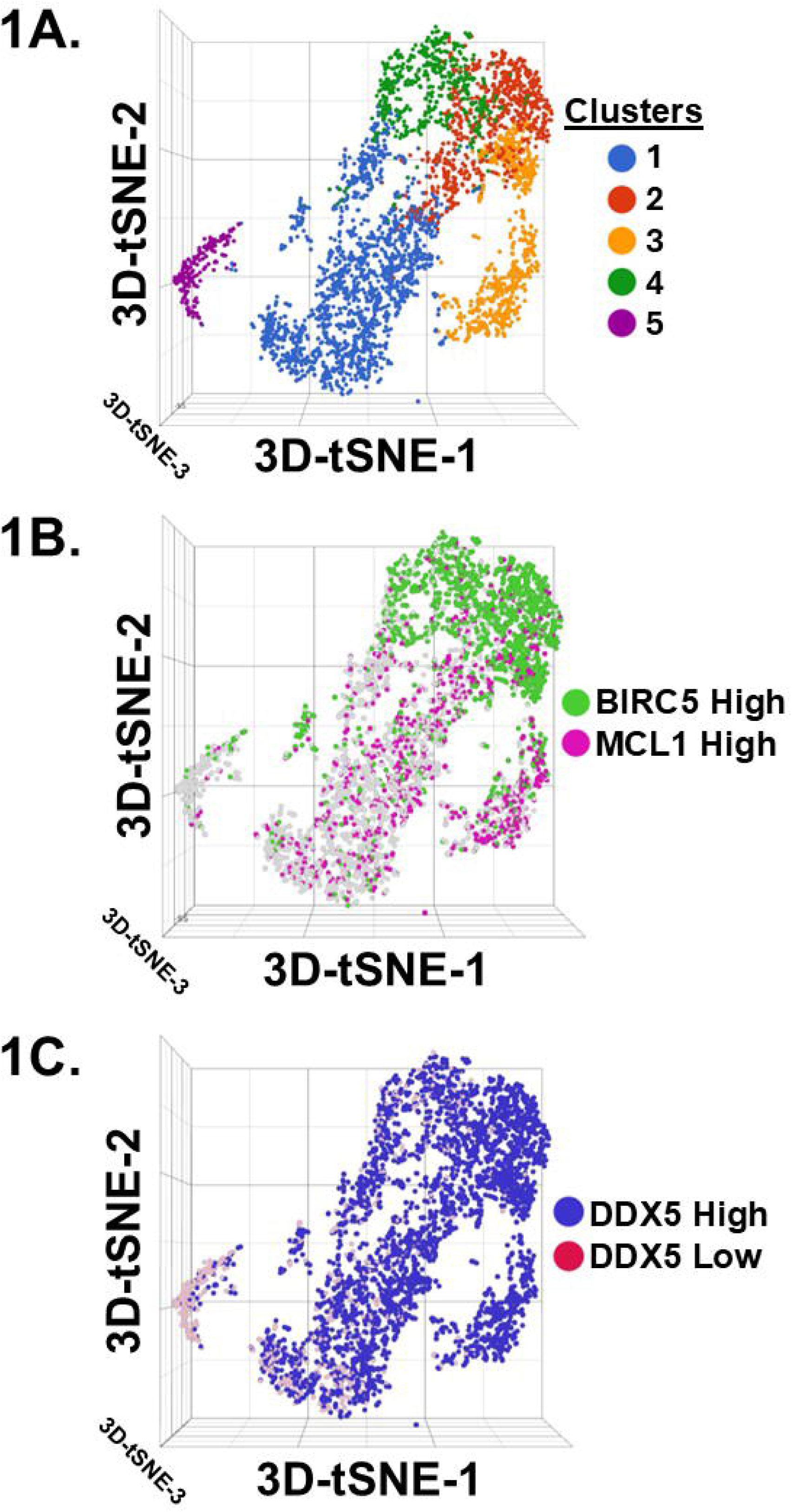
Baseline single-cell RNAseq analysis on the MCL cell line pair Mino P/R (n=3841 cells). **1A)** Comparison of the t-SNE/Graph-based clusters between MINO P vs MINO R cell lines. **B-C)** Figures showing single cells with enriched expression of the target genes BIRC5, MCL-1, and DDX5.

### SecDrug treatment inhibits human MCL cell line proliferation as a single agent and synergizes with Proteasome inhibitors and BTK inhibitors

We first evaluated the *in vitro* cytotoxic effect of YM155 and S63845 as single agents against MCL cell lines representing drug-sensitive (JEKO1, MINO-P), Proteasome inhibitor-resistance (MINO-VR), and BTKi-resistance (Z-138) as *in vitro* model systems. We found that both YM155 and S63845 effectively reduced cell viability in all four MCL cell lines, irrespective of PI/BTKi sensitivity/resistance (**Figure 2A-B**). The half-maximal inhibitory concentration (IC_50_) range of single-agent YM155 and S63845 in human MCL cell lines were 1.57-5.64nM and 250nM-4.7µM, respectively.

**Figure 2.**
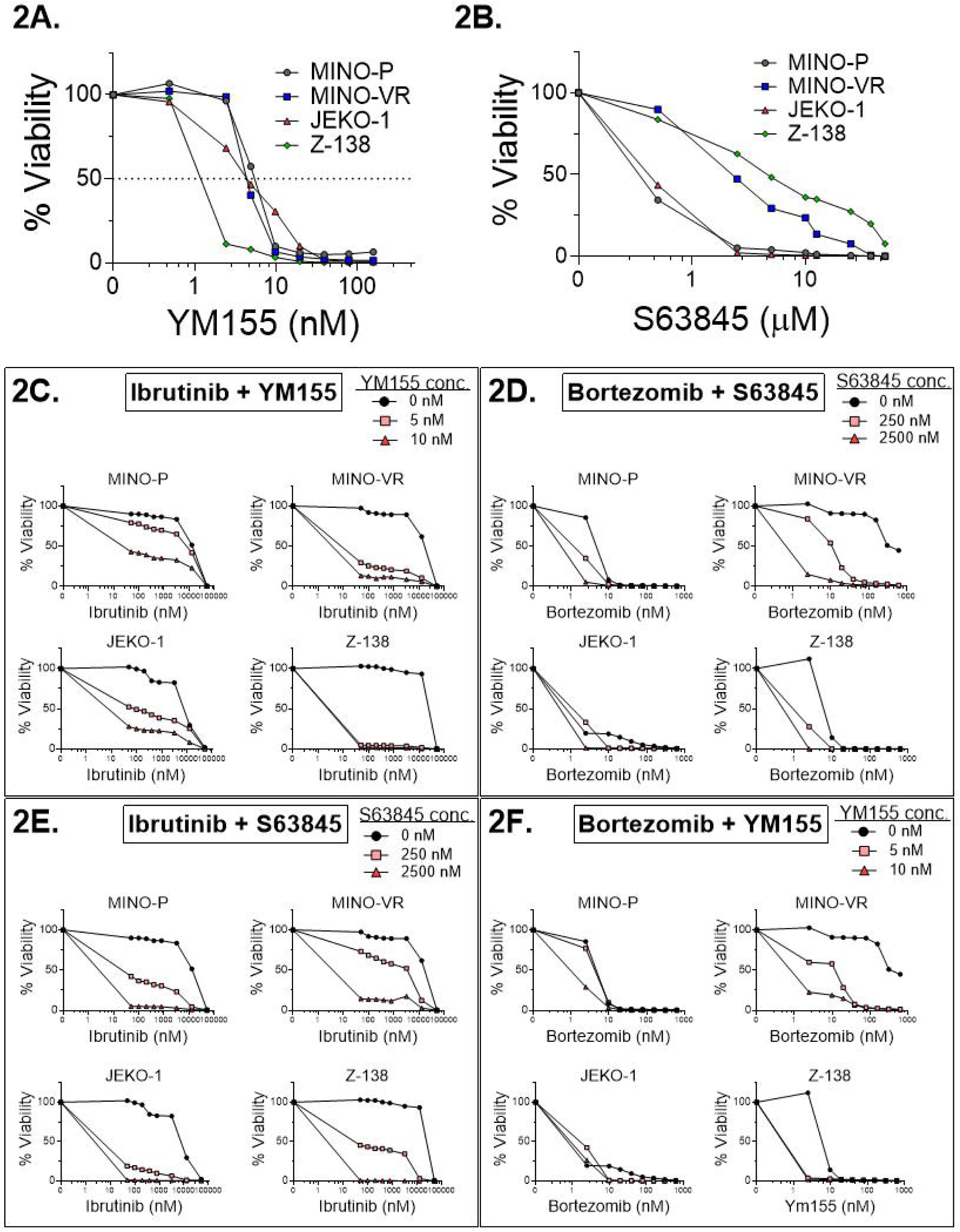
*In vitro* cell viability assays. Dose-response plots in MCL cell lines showing single-agent *in vitro* cytotoxicity of **A)** YM155 and **B)** S63845. Both YM155 & S63845 showed high single-agent *in vitro* cytotoxicity in our MCL cell panel, including PI-resistant and BTKi-resistant MCL cell lines. **C-F. YM155 & S63845 exhibit synergistic cell-killing activity when combined with PI and BTKi.** Dose-response plots represent the *in vitro* cell viability profile of MCL cell lines, including PI-resistant and BTKi-resistant MCL cell lines treated with different combinations of **C-D**) YM155 and **E-F**) S63845 with PI and BTKi. The combination index (CI) and Dose reduction index (DRI) values were calculated by Calcusyn Software (BioSoft) using Chou-Talalay’s algorithm. All the combinations showed significant improvement in lowering cellular proliferation as compared to the effect of PI/ BTKi alone, which indicates drug synergy (CI<0.9). Synergistic effects were particularly profound in MINO-VR and Z-138. This observation is particularly relevant as these lines represent acquired and innate resistance, respectively.

Next, we investigated the impact of different concentrations of YM155 and S63845 in combination with an increasing dose range of Bortezomib (PI) or Ibrutinib (BTKi). We observed that all the combination treatment regimens showed higher cytotoxic effects than single-agent PI or BTKi treatment (**Figure 2C-F**). YM155 and S63845 exhibited synergistic inhibitory activities measured as combination index (CI) values calculated using Chou-Talalay’s CI theorem (CI<0.9 depicts synergism). Most combinations showed synergy, and synergistic effects were particularly profound in the R/R MCL cell lines MINO-VR and Z-138. This observation is particularly significant since these cell lines represent resistance to PIs and BTKi, respectively. Further, our results also showed that both YM155 and S63845, in combination with BTKi/PI, were able to lower the effective dose of both BTKi and PI significantly required to achieve the desired therapeutic response by > 12 times. The mean Dose Reduction Index (DRI) for YM155 combination therapy is 15.87 ± 4.93, and for S63845 combination therapy is 12.34 ± 2.67, providing evidence supporting a sensitizing effect.

### Combination of secDrugs (BIRC5-inhibitor + MCL1-inhibitor) has a synergistic effect

Next, using *in vitro* cytotoxicity assay, we observed that these two secDrugs (YM155+S63845) have substantial synergistic cell-killing action in MCL cells (**Figure 3A**). Drug synergy scores were then calculated using Chou-Talalay’s Combination Index theorem, as described above. We observed that the constant ratio combination of YM155 and S63845 shows significant synergy (CI value at Fraction affected=0.5 was 0.550, while the DRI for YM155 and S63845 were 2.454 and 7.032, respectively; **Figure S1**), indicating that the combination of these two secDrugs will further improve the cancer-killing effect.

**Figure 3.**
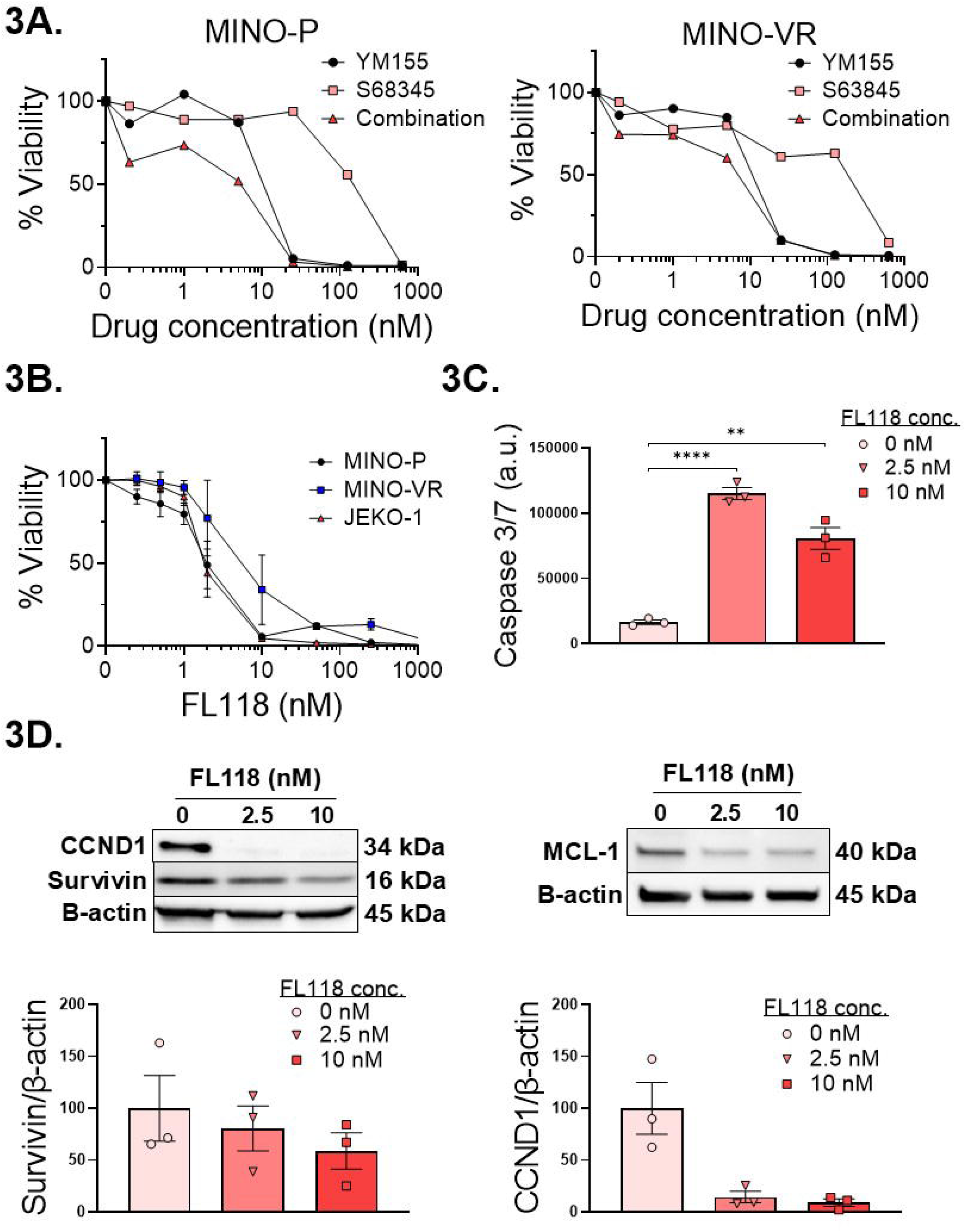
Combination of secDrugs (BIRC5-inhibitor + MCL-1-inhibitor) has a synergistic effect. **A)** Representative plots showing secDrug-secDrug (YM155 + S63845) combination therapy analysis in sensitive and resistant MCL cell lines. Results show that the YM155-S63845 combination has synergistic activity (CI<0.9). **B)** *In vitro* cytotoxicity of FL118 in MCL cell lines. FL118 is a small-molecule inhibitor that potentially downregulates both MCL-1 and BIRC5. **C)** FL118 treatment induces caspase 3/7 activity at 24h. **D)** FL118 target validation. MCL cells treated with FL118 show decrease in BIRC5, S63845, and CCND1 protein expression.

FL118 is a structural camptothecin-related small molecule that potentially downregulates both MCL1 and BIRC5^23,24^. The representative plots in **Figures 3 B-D** show that MCL cells treated with FL118 have notable *in vitro* cytotoxicity (IC_50_ <10nM), decreased BIRC5, MCL1, and CCND1 protein expression, and induced caspase 3/7-mediated apoptotic activity.

### secDrugs augment apoptosis in sensitive and resistant MCL cells via mitochondrial-mediated pathway

To confirm whether the loss of cell viability was indeed due to apoptosis, quantitative analysis of the extent of apoptosis in MCL cells in response to secDrug single-agent treatment and in combination with PI/BTKi was done using Fluorescein isothiocyanate (FITC) conjugated Annexin-V staining, followed by flow cytometry (**Figure 4A**). We observed a higher population of Annexin-V positive cells upon treatment with secDrugs compared to no treatment, indicating apoptosis. Drug-induced apoptosis of YM155 and S63845 and the synergistic activity of the drug combination were confirmed by Caspase 3/7 activity (**Figure 4B**).

**Figure 4.**
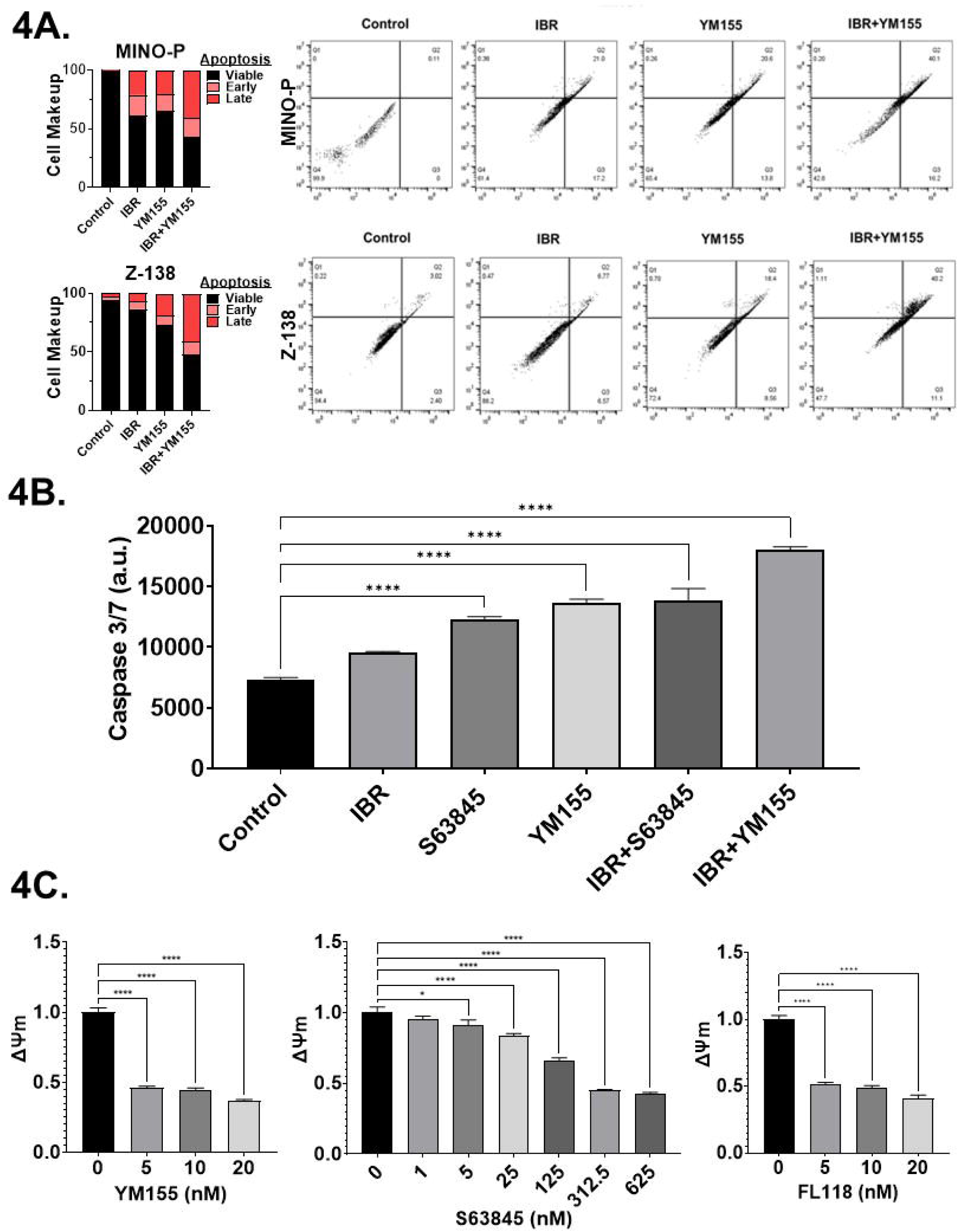
Apoptosis assays. **A)** Apoptotic cells representing the effect of secDrug on MCL cell lines as single agent and in combination with BTKi. Cells were treated for 48 hours, and then apoptosis was measured using Annexin V-FITC and flow cytometry. Results show a significantly higher population of cells that are Annexin V positive in combination treatment as compared to single agent treatment, indicating an elevated level of induction of apoptosis. **B)** Representative Caspase 3/7 activity assays showing increased apoptosis following YM155 and S63845 single-agent and combination treatment. **C)** secDrug treatment affects mitochondrial membrane potential (MMP) in MCL cells. Representative plots confirm the decrease in mitochondrial membrane potential following YM155, S63845, and FL118 treatment. JC-1 (Sigma), a cationic carbocyanine dye that accumulates in healthy mitochondria and forms aggregates, was used to assess Mitochondrial membrane potential (ΔΨ_M_).

Further, secDrug-dependent mitochondrial dysfunction was confirmed by measuring the mitochondrial membrane potential using JC-1 dye. JC-1 is a cationic carbocyanine dye that accumulates in the matrix of healthy mitochondria and forms J-aggregates, which emit red fluorescence, while its accumulation decreases in depolarized Mitochondria of apoptotic cells, and JC-1 remains in a monomeric form, which emits green fluorescence. Thus, a decrease in the ratio of red/green fluorescence indicates mitochondrial depolarization. We observed a significant shift from red to green fluorescence in response to YM155 and S63845 treatment, as well as with FL118, indicating mitochondrial dysfunction/depolarization due to loss of mitochondrial membrane potential (ΔΨ) (**Figure 4C**).

### YM155 targets cancer stem cells in MCL and reduces ALDH activity

As YM155 has reported activity against cancer stemness^25^, we further investigated the effect of our novel drugs on the cancer stem cells (CSCs) in MCL, which have a potential role in treatment resistance^26^. Aldehyde dehydrogenase (ALDH) is an intracellular detoxification enzyme frequently overexpressed in CSCs and involved in drug resistance. We observed >10-fold higher ALDH activity in the drug-resistant MCL lines compared to the drug-sensitive MCL lines (**Figure 5A**). Further, YM155 single-agent treatment led to >90% reduction in ALDH activity in resistant MCL cells compared to the untreated control (**Figure 5B**). This is consistent with the previous finding that BIRC5 expression overlays with stem cell markers^25^.

**Figure 5.**
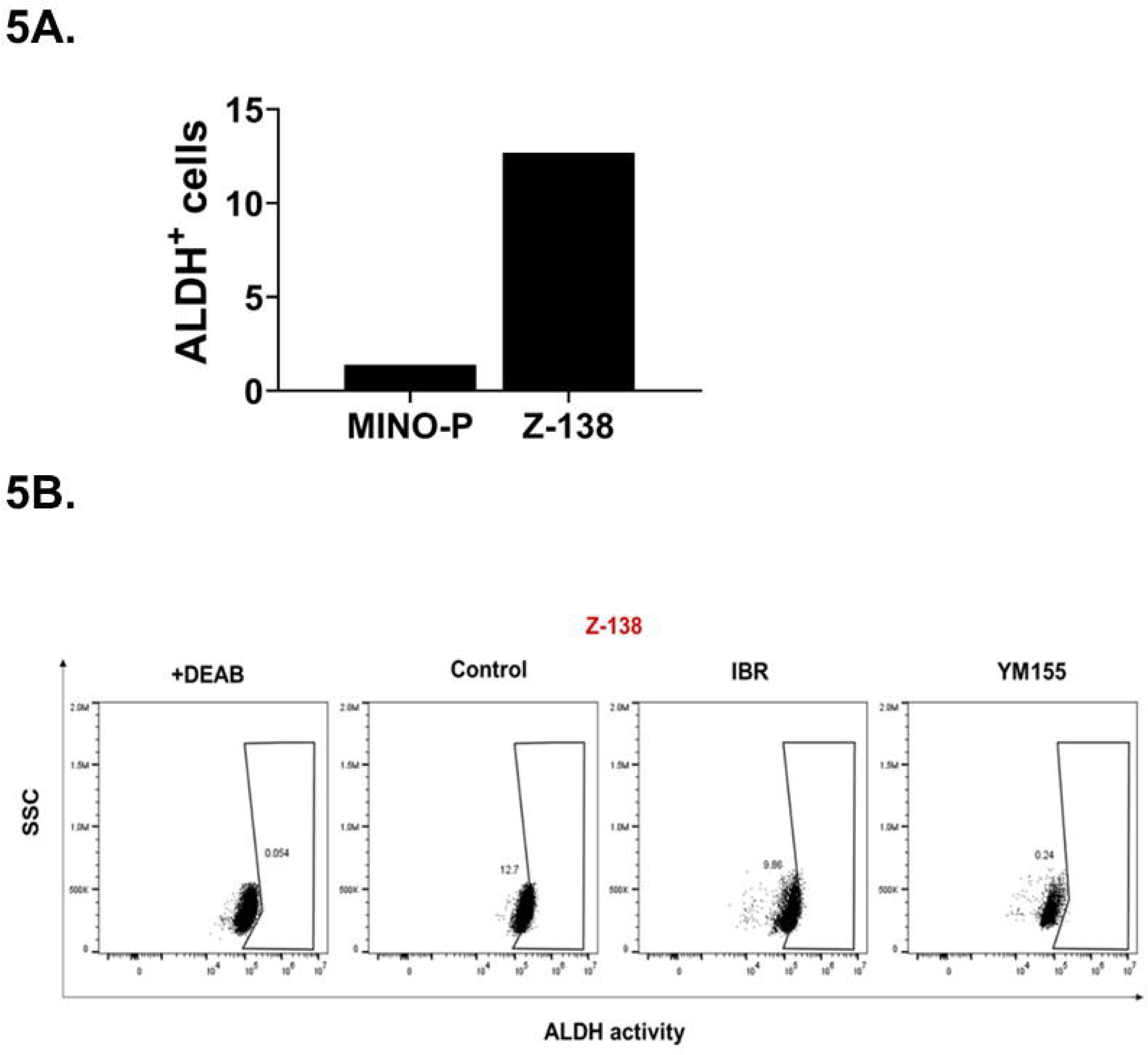
Aldehyde dehydrogenase (ALDH) activity in MCL cell lines following YM155 treatment. **A)** Very low ALDH activity was observed in the sensitive cell line MINO-P compared to the innate resistant Z-138 cell line. **B)** YM155 treatment led to a significant decrease in ALDH activity (>90%) in drug-resistant cells as compared to single-agent Ibrutinib treatment (∼29%). N,N-diethylaminobenzaldehyde (DEAB) - a reversible, competitive inhibitor of ALDH, was used as a positive control.

### Gene expression profiling revealed genes and molecular pathways/networks associated with the mechanism of action of secDrugs in MCL cells

Heatmaps and Venn Diagrams in **Figure 6A-B** represent the results of differential gene expression analysis of secDrug treatment-induced changes (untreated vs. single-agent treatment) in MCL cell lines using a modified Limma method called Gene Specific Analysis (details in the Methods section).

**Figure 6.**
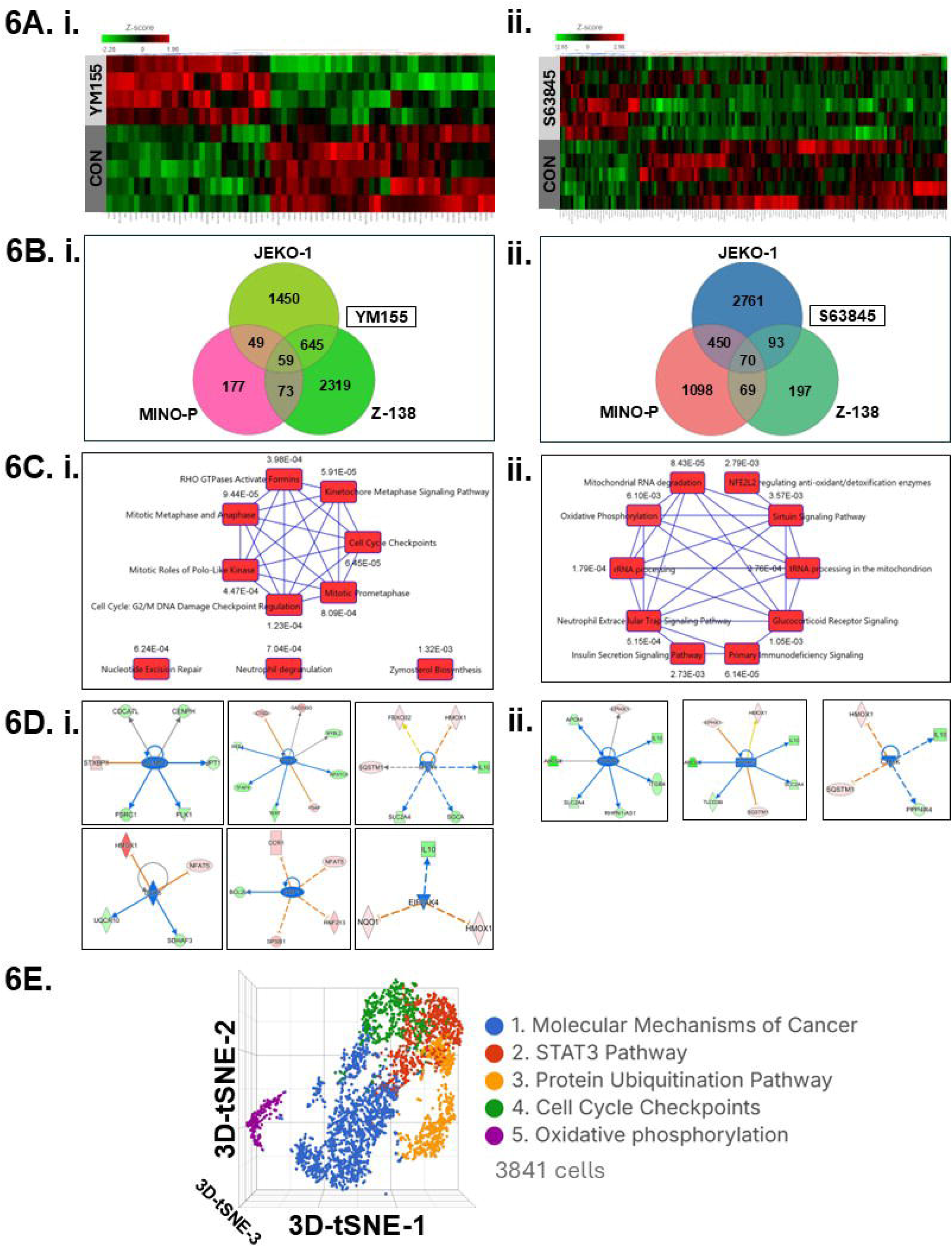
Gene expression analysis. **A.** Heatmaps representing the top genes that were most significantly differentially expressed in MCL cell lines. **B.** Venn Diagrams showing treatment (YM155, BTZ, and IBR) regulated genes that were common between the cell lines. **C-D.** IPA analysis predicted secDrug-induced key canonical pathways and Upstream regulators. i**) YM155 ii) S63845. E.** IPA analysis of single-cell clusters showing enrichment of top pathways for each cluster.

A total of 1221 genes were significantly differentially expressed (DEGs) (p<0.05; fold-difference ≠ 1) following YM155 treatment. Among these, 690 genes had a |fold-change| ≥ 1.5 (p<0.05; **Figure 6A.i**). When cell line-wise DE analysis (Treated vs. Untreated) was performed, 194 DEGs were shared at |fold-change| > 1.5 (Venn diagram; **Figure 6A.ii)**. As shown in **Figure 6 B.i,** the differential gene expression signature of **S63845** consists of 367 genes (p<0.05; fold-difference ≠ 1). 131 genes had a |fold-change| ≥ 1.5. The Venn diagram in **Figure 6B.ii** shows that 70 DE genes were shared between the MCL cell lines. The top 50 DEGs for each single-agent treatment (YM155 and S63485) are listed in **Table 1 A-B**. The following 5 DEGs were common between YM155 and S63845 single-agent treatments: ABCG1, EPHX1, SQSTM1, GADD45G, and NQO1. Interestingly, the top genes included in the single-agent DEG signatures showed further dysregulation following combination treatment, implying a synergistic effect on mRNA expression. **Table S2 A-D** shows the top DEGs for each treatment combination in the sensitive line MinoP and the BTZ-resistant line MinoVR: YM155+BTZ, YM155+IBR, S63845+BTZ, and S63845+IBR.

Ingenuity pathway analysis revealed regulation of mitosis & cell cycle checkpoints and mTOR signaling as the top canonical pathways in YM155-treated MCL cell lines (**Figure 6C**), while mitochondrial dysfunction and oxidative phosphorylation were the top pathways for S63845. Based on the list of top DEGs, IPA predicted downregulation of the following upstream regulators following secDrug treatment: DDX5, CCND1, Ikaros (IZKF1), EIF2AK4, PPARG, and FOXA1 as top upstream regulators (**Figure 6D**). The identification of DDX5 as an upstream regulator is highly relevant since our recent studies have indicated that both BIRC5 and MCL1 are downstream targets for DDX5^27^.

Interestingly, IPA analysis of genes up-regulated in single cells showed enrichment of several of the top secDrug-induced dysregulated pathways in the t-SNE clusters representing baseline expression of MCL cell lines (**Figure 6E**).

### Target Validation

Next, we validated the on-target effect of YM155 in MCL cells using western blotting. Representative blots in **Figure 7A** show that YM155 down-regulates the expression of BIRC5 protein in MCL cells, validating the target-specific effect of this secDrug to exert its anti-cancer effect. Further, we show that YM155 significantly downregulates BIRC5 mRNA expression (**Figure 7B**).

**Figure 7.**
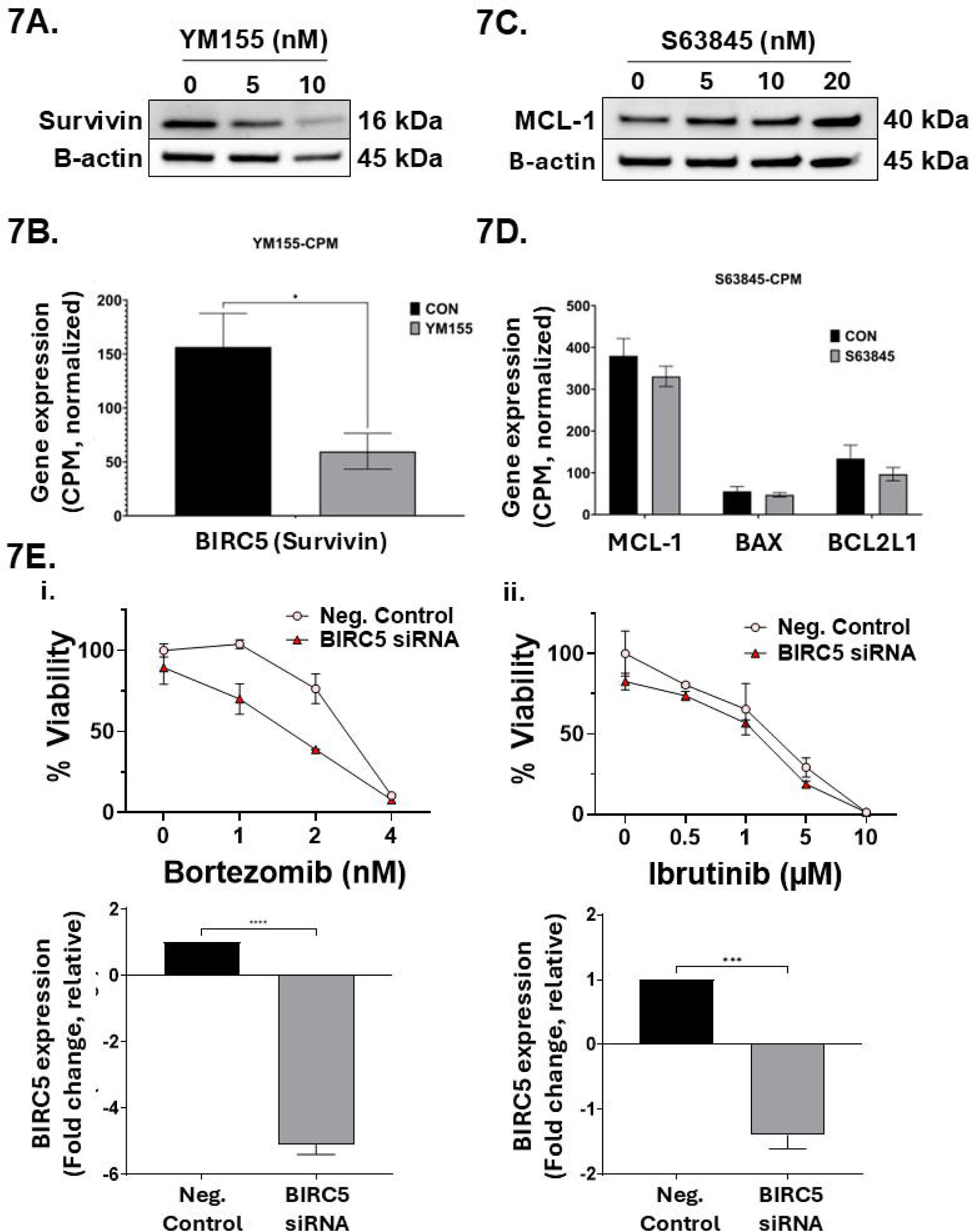
Target validation. Representative western blots show **A)** YM155 significantly down-regulates Survivin/BIRC5 protein, but **C**) S63845 increases MCL-1 expression. **B)** YM155 significantly downregulates BIRC5 mRNA expression, while **D)** S63845 has minimal effect on MCL-1, BAX, and BCL-xL mRNA levels in MCL cells. **E.** siRNA knockdown of BIRC5 gene expression shows higher *in vitro* cytotoxicity following i) BTZ and ii) Ibrutinib treatment at 24h compared to wild-type cells. Results of the corresponding qPCR analysis of BIRC5 mRNA (n = 3) are also provided.

Interestingly, treatment of MCL cells with S63845 increased the protein expression of its target, MCL1, but not MCL1 mRNA (**Figure 7C, D**). This has also been observed earlier, where S63845 treatment-induced increase in MCL1 protein level was shown to be correlated with protein half-life^28^. Further, we also concur that the BH3 mimetic S63845 had minimal effect on BCL-2-and BCL-XL mRNA expression^29^ (**Figure 7D**).

Additionally, RNA interference was used in MCL lines to confirm the effect of inhibition of the targets of the top secondary drug candidate predicted by our secDrug algorithm. Using siRNA-based knockdown, we inhibited BIRC5 in the MINO-P cell line (qPCR results are shown in **Figure 7E**) and evaluated the relative cell viability via 24hr dose-response study following treatment with PIs and BTKis. We observed a modest decrease (∼15%) in cell viability in cells transfected with target-specific siRNA compared to our negative transfection control. When these BIRC5-knockdown cells were treated with Bortezomib or Ibrutinib, increase in the *in vitro* cytotoxicity was observed compared to our drug-treated negative controls (wild type) (**Figure 7E**).

### Validation of top differentially expressed genes in patient datasets

Next, we used gene expression data (GSE141335) on primary samples from patients with MCL and compared the association of top treatment-induced gene signatures with *ex vivo* IBR response. Our results show that the expression of several genes was dysregulated by YM155 (**Figure 8A.i**) and S63845 (**Figure 8A.ii**) treatment, which are also significantly associated with IBR sensitivity, indicating that these secDrugs are capable of reversing the oncogenic progression and drug resistance in MCL.

**Figure 8.**
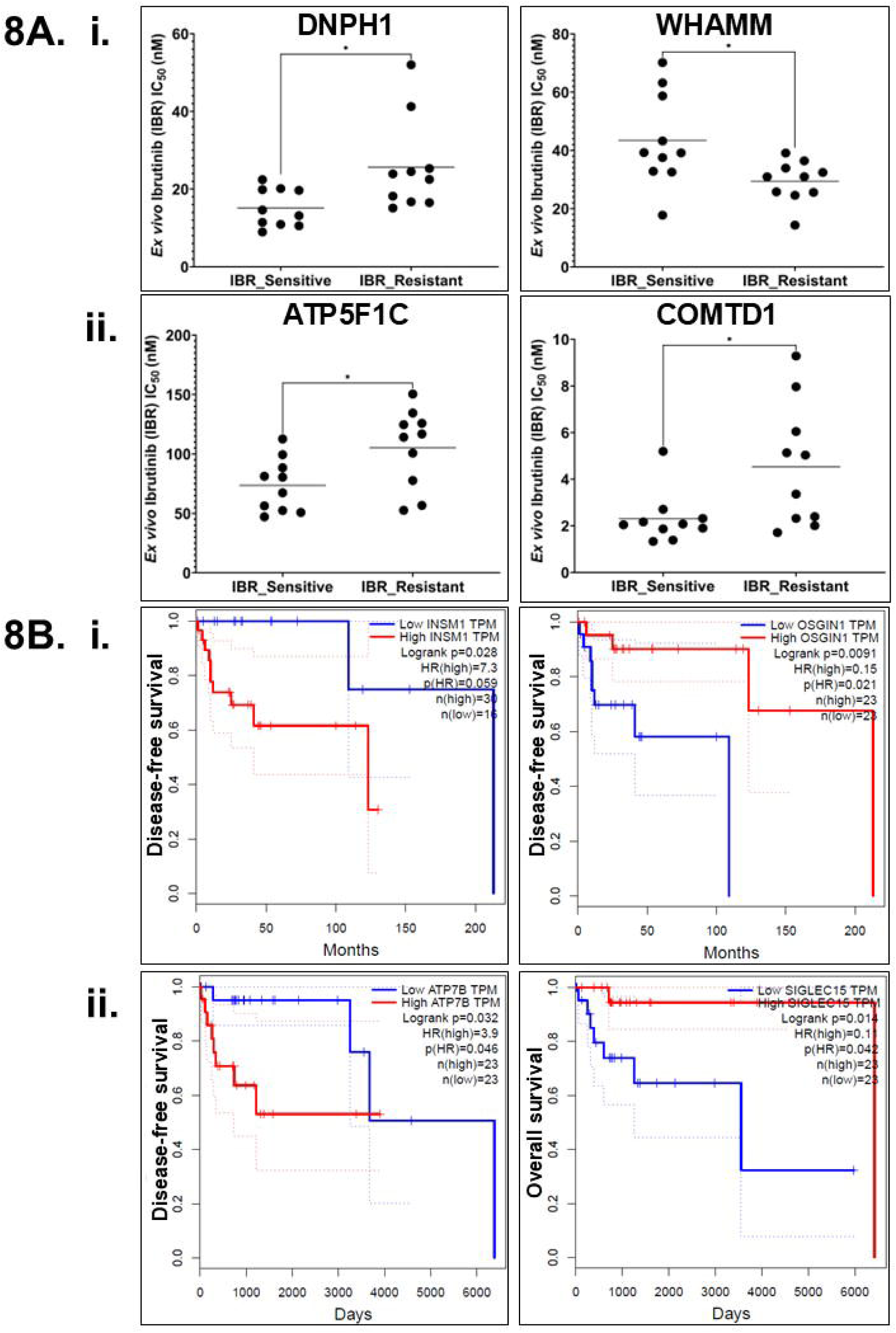
*In silico* validation using secDrug treatment signatures. **A)** *Ex vivo* IBR sensitivity plots in primary samples from MCL patients, and **B)** Kaplan-Meier plots showing secDrug treatment-associated genes are significantly associated with survival in lymphoma patients from the TCGA database. i) YM155 gene signatures. ii) S63845 gene signatures.

Finally, we performed *in silico* validation of the differentially expressed genes associated with S63845 and YM155 treatment using The Cancer Genome Atlas’s (TCGA) Diffuse Large B-cell lymphoma (DLBC) gene expression profiling dataset. Kaplan-Meier Curves showed that several of the top DE genes for YM155 (**Figure 8B.i**) and S63845 (**Figure 8B.ii**) were significantly associated with clinical outcomes, i.e., overall survival and disease-free survival.

## DISCUSSION

Most patients with MCL have advanced (stage III or stage IV) disease at diagnosis, with a progression-free survival of ∼1-2 years and median overall survival (OS) of <3 years^30,31^. BTKi (Bruton’s tyrosine kinase inhibitors), such as Imbruvica (ibrutinib), Calquence (acalabrutinib), and Brukinsa (zanubrutinib), are standard targeted therapeutic options for refractory or relapsed (R/R) MCL^32^. BTK is a non-receptor tyrosine kinase that serves as a key component of the B-cell receptor signaling pathway responsible for cellular proliferation and is found to be frequently overexpressed in MCL^33,34^. Further, the proteasome inhibitor (PI) drug Bortezomib (Velcade/BTZ) is another FDA-approved targeted drug for R/R MCL^35,36^. However, despite these advances in the treatment landscape, MCL remains incurable with limited therapeutic options owing to drug resistance, extensive inter-individual variation in response, and toxicity profile that limits efficacy in clinical settings^9,37,38^. Once ibrutinib stops working, only one-third of patients respond to the next line of treatment, and those who do respond experience only brief remissions and have poor outcomes, irrespective of stem cell transplantation^39^ Therefore, there is an unmet need to discover novel drugs against R/R MCL.

Our novel computational algorithm, secDrug, identified BIRC5 and MCL1 as potential secondary targets for the treatment of R/R MCL. BIRC5 is a member of the inhibitor of apoptosis (IAP) protein family that inhibits apoptosis through caspase-dependent and independent pathways, blocks cell death, and influences resistance to anti-cancer therapies^40,41^. BIRC5 is found in approximately 50% of patients with high-grade non-Hodgkin’s lymphomas, where it is an adverse prognostic factor since high BIRC5 expression is correlated with shorter survival^42,43^. Earlier studies have shown that MCL tumor cells have 17q gain, leading to the upregulation of BIRC5 expression^44^. MCL1 is a pro-survival protein that regulates the mitochondrial apoptotic pathway. MCL1 belongs to the BCL-2 family of proteins that are characterized by the presence of BCL-2 homology (BH) domains^28^. Other members of this group include BCL-2, BCL-xL/ BCL2L1, BCL-W, MCL1 and BFL1/BCL2A1. Upregulation of MCL1 has been implicated in the growth of tumors in several cancers, including multiple myeloma, pre-B and B lymphomas, while the downregulation of MCL1 may lead to cytotoxicity in tumor cells^45–48^.

We confirmed our secDrug predictions of BIRC5 and MCL1 using a novel single-cell transcriptomics-based target gene screening approach in Mantle Cell Lymphoma cell lines. To validate these findings using cell-based assays, we used the small molecule inhibitors of Survivin/BIRC5 (YM155) and MCL1 (S63845) to demonstrate that these genes are promising therapeutic targets in the management of R/R MCL. Earlier studies in several other hematological malignancies have suggested that both YM155 and S63845 are potent in targeting drug-resistant sub-clones^49–51^.

Here, we showed that inhibition of BIRC5 and MCL1 exhibited significant synergistic cell-killing activities alone and in combination with Bortezomib (PI) and Ibrutinib (BTKi), especially in R/R MCL cells. An earlier study screened ∼3,800 clinically approved drugs and drug candidates in JEKO1 and MinoP and identified 98 compounds, including YM155, alisertib (Aurora kinase inhibitor), and carfilzomib (a proteasome inhibitor), with >50% inhibition in either MCL cell line^52^. S63845 is a selective MCL1 inhibitor that induces significant alterations in mitochondrial function and stress-response genes^53^. Notably, we also show the benefit of combining these two secDrugs (YM155 and S63845) as an effective dual inhibition strategy against MCL. Further, since BIRC5 and MCL1 are downstream of DDX5, and our IPA analysis also predicted downregulation of the upstream regulator DDX5 as a potential consequence of treatment with these secDrugs, we hypothesized that the small molecule inhibitor of DDX5, FL118, has anti-cancer benefits in aggressive MCL through the simultaneous inhibition of these downstream targets. This hypothesis is consistent with the previous findings ^23,54^. Early studies indicated that FL118, a camptothecin-related small molecule, downregulates key cancer cell survival proteins, including MCL1, cIAP, BIRC5, and XIAP^27^. Indeed, we showed that treatment with FL118 has high levels of efficacy in R/R MCL and downregulates both BIRC5 and MCL1 protein expression. We plan to further validate the effectiveness of dual inhibition of BIRC5 and MCL1 as a therapeutic strategy against R/R MCL using our novel fluoroaryl-substituted derivatives of FL118, including FL77-9, FL77-24, and FL77-32 (7h), which have earlier shown significant *in vitro* cytotoxicity and *in vivo* antitumor efficacy in solid tumor models^23,27,55^.

ALDH expression has been shown to be associated with cancer stem cells (CSCs) and aggressiveness in several cancers^56^. ALDH+ MCL cells are found to be highly clonogenic, relatively quiescent, and resistant to chemotherapy^56,57^. We observed that YM155 was remarkably effective in reducing ALDH activity in resistant MCL cells.

Next, we performed pre-vs-post treatment next-generation RNA sequencing analysis to identify mechanisms of secDrug action and synergy. Gene expression profiling and pathway analysis of YM155-treated MCL cell lines revealed down-regulation of the pro-survival pathway and mTOR pathway genes, as well as up-regulation of pro-apoptotic markers and several pathways associated with cancer aggressiveness. Further, YM155 disrupts the expression of several genes linked to mitosis, chromosomal stability, and cell survival in mantle cell lymphoma. These include key mitotic regulators AURKB (Aurora kinase), NDC80, SPC24, CENPV, and CENPH, which are downregulated, impairing the chromosomal passenger complex and kinetochore-microtubule interactions, potentially driving apoptosis and mitotic errors^58–60^. Overexpression of these genes is associated with poor prognosis or oncogenic progression in various cancers^58–60^. We also observed YM155-induced downregulation of PIMREG, which may disrupt cell cycle progression^61^.

S63845 treatment resulted in significant upregulation of several mitochondrial genes, MT-CYB, MTND-2, and MTNDP424, suggesting a compensatory response to mitochondrial stress induced by MCL1 inhibition, as MCL1 is known to affect mitochondrial integrity^62,63^. An increase in SQSTM1 and FBXO32 is indicative of enhanced autophagy and proteasomal degradation, indicating tumor suppressor effects^64,65^. We also observed increases in the ferroptosis-related genes (HMOX1, SNORD3A), which are likely up-regulated as cellular defense against DNA damage and oxidative stress^66,67^. Interestingly, NQO1 and GADD45G, stress-response genes with tumor-suppressing functionality and apoptosis induction to cellular stress, were up-regulated upon both YM155 and S63845 treatments^66^. Several of the top DEGs associated with YM155 or S63845 single-agent treatments were further dysregulated following combination treatments with BTZ or IBR, indicating possible mechanisms of drug synergy, which we plan to investigate further. Finally, *in silico* analysis of our gene signatures with GEP datasets from patients suggested that secDrug treatment was capable of reversing dysregulated signatures associated with tumor survival, cell proliferation, differentiation, apoptosis (DNPH1), oxidative phosphorylation, mitochondria-regulated pathways, cancer progression, drug resistance, metastasis (ATP7B, ATP5F1C; COMTD1), and autophagy (WHAMM)^68–71^.

In summary, our study underlines the importance of BIRC5 and MCL1 as novel therapeutic targets in aggressive MCL models and provides additional novel drugs to overcome resistance to standard-of-care drugs. Our work lays the groundwork for additional pre-clinical investigation to establish the inhibition of these novel mechanisms for the treatment of aggressive forms of MCL.

## Data Availability

The datasets generated during and/or analyzed during the current study are available from the corresponding author upon reasonable request.

## ACKNOWLEDGEMENTS

The authors would like to thank Prof. Brian Van Ness for his valuable feedback and insightful contributions to the manuscript. S.M. was a postdoctoral researcher at Auburn University when this work was completed and is currently employed at Champions Oncology, Inc.

## AUTHOR CONTRIBUTIONS

Conceptualization: A.K.M.

Methodology, Investigation & Validation: J.P., S.C., and S.M;

Data Analysis: J.P., S.C., U.K.M., and A.K.M;

Visualization: J.P., S.C., and A.K.M;

Writing: original draft: J.P., S.C., and A.K.M;

Writing: review & editing: F.L. and A.K.M;

Refocus: F.L.;

Supervision & Project administration: A.K.M.

All authors have read and agreed to the published version of the manuscript.

