## Supplementary Material for "Inhibition of BIRC5 and MCL1 as a potential treatment strategy to overcome drug resistance in Mantle Cell Lymphoma"

**This file includes:**

Supplementary Text

Figs. S1 to S2

Tables S1 to S2

Fig. S1.

**Representative isobologram showing secDrug-secDrug (YM155 + S63845) combination therapy in MCL cell line.** Results show that the YM155+S63845 combination has synergistic activity (CI<0.9; DRI>1).

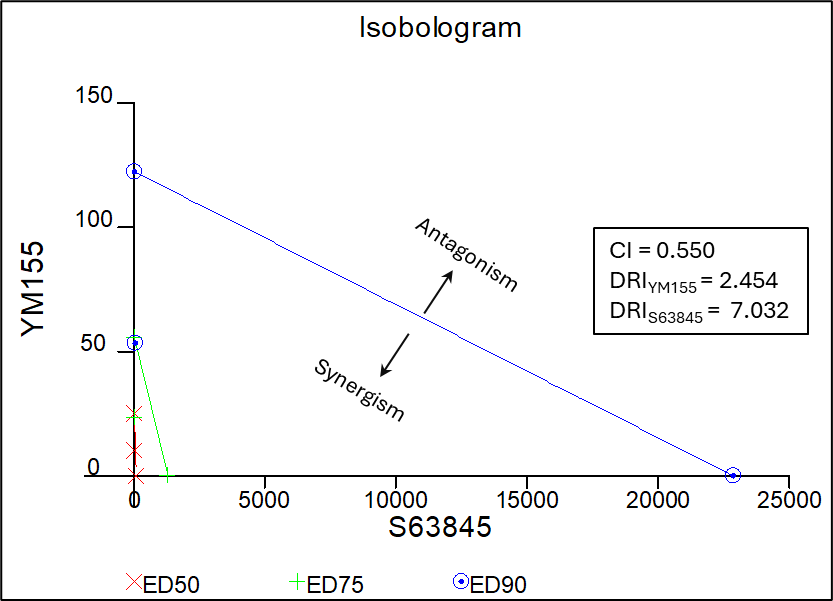

Fig. S1.

**Single-agent cytotoxicity assay with proteasome inhibitor (PI) & Bruton’s Tyrosine kinase inhibitor (BTKi) shows extensive inter-individual variation in drug response in a MCL cell line panel.** Dose-response curves reveal a wide range of drug sensitivity towards (Figure 1A) the PI Bortezomib (BTZ) and (Figure 1B) the BTKi Ibrutinib (IBR). MINO-VR has an approximate 70 folds higher IC_50_ value for Bortezomib than its parental cell line MINO-P. Z-138 has an approximate 2.5 folds higher IC_50_ value for Ibrutinib than MINO-P and JEKO-1.

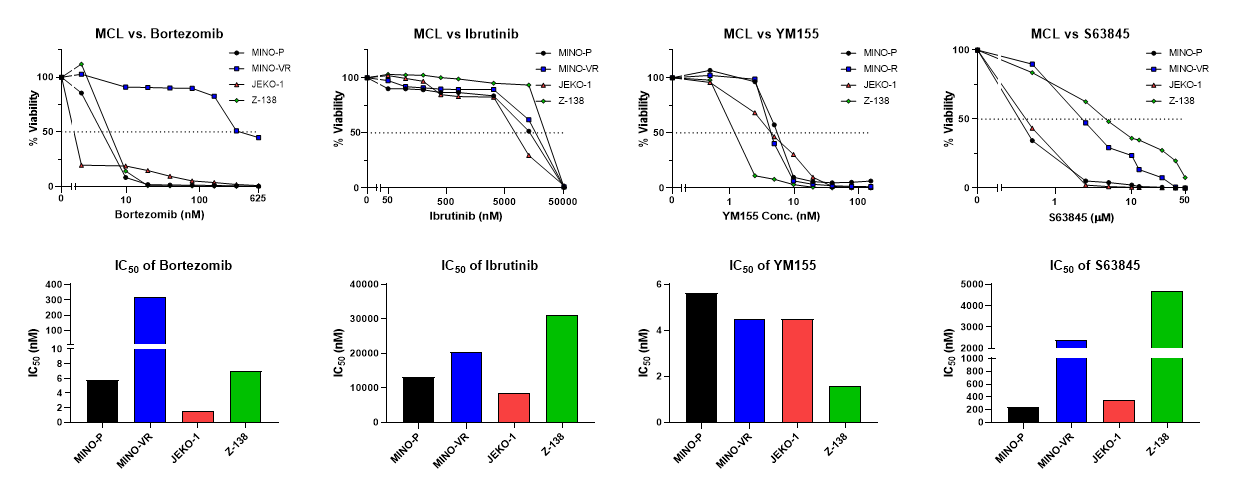

**Table S1.** **List of drugs, reagents, antibodies, and kits.**

| **Reagents** | **Manufacturer** | **Location** |
| --- | --- | --- |
| **Fetal bovine serum (FBS)** | Hyclone (Thermo-Fisher Scientific Inc.) | Rockford, IL, USA |
| **Trypsin (0.25% w/v)** | Hyclone (Thermo-Fisher Scientific Inc.) | Rockford, IL, USA |
| **Recombinant Human IL-6** | PeproTech, Inc. | Cranbury, NJ, US |
| **Penicillin-Streptomycin (10,000 U/mL)** | Gibco^TM^ (Thermo-Fisher Scientific Inc.) | Waltham, MA, USA |
| **FITC Annexin V Apoptosis Detection Kit** | BD Biosciences | San Jose, CA, USA |
| **RIPA lysis buffer** | Thermo-Fisher Scientific Inc | Waltham, MA, USA |
| **Halt™ Protease and Phosphatase Inhibitor Cocktail** | Thermo-Fisher Scientific Inc | Waltham, MA, USA |
| **Pierce™ ECL Western Blotting Substrate** | Thermo-Fisher Scientific Inc | Waltham, MA, USA |
| **RNeasy Plus Mini Kit** | QIAGEN | Hilden, Germany |
| **Quick Start Bovine Serum Albumin Standard** | Bio-Rad | Hercules, CA, USA |
| **Tris Buffer Saline (TBS)** | Bio-Rad | Hercules, CA, USA |
| **10% Tween 20** | Bio-Rad | Hercules, CA, USA |
| **Polyvinylidene fluoride membrane (PVDF)** | EMD Millipore | Billerica, MA, USA |
| **Bovine Serum Albumin (BSA)** | VWR | Radnor, PA, USA |
| **Dimethyl sulfoxide (DMSO)** | Sigma-Aldrich Inc | St. Louis, MO, USA |
| **Bradford Reagent** | Sigma-Aldrich Inc | St. Louis, MO, USA |
| **JC-1 - Mitochondrial Membrane Potential Assay Kit** | Abcam | Waltham, MA, USA |
| **Anti-rabbit IgG, HRP-linked Antibody (7074S)** | Cell Signaling Technology | Danvers, MA, USA |
| **β-actin (A3854)** | Sigma-Aldrich Inc | St. Louis, MO, USA |
| **Calcein AM, cell-permeant green and blue dyes** | Thermo-Fisher Scientific Inc | Waltham, MA, USA |
| **S63845** | Selleckchem | Houston, TX, USA |
| **YM155** | Selleckchem | Houston, TX, USA |
| **MCL** | Cell Signaling Technology | Danvers, MA, USA |
| **Survivin** | Cell Signaling Technology | Danvers, MA, USA |

Table S2. Top DEGs for each treatment combination in the sensitive line MinoP (A. YM155+IBR, B. S63845+IBR) and the BTZ-resistant line MinoVR: (C. BTZ+YM155, D. BTZ +S63845).

S2.A. IBR+YM155 Combination

| **Gene name** | **P-value (MINO-P,IBR_YM155 vs MINO-P,CON)** | **Fold change (MINO-P,IBR_YM155 vs MINO-P,CON)** |
| --- | --- | --- |
| **CD72** | 1.74E-02 | **-15.21** |
| **TERT** | 6.54E-05 | **-14.26** |
| **SPIB** | 3.98E-02 | **-12.35** |
| **ABCG1** | 1.82E-03 | **-11.91** |
| **SIT1** | 2.21E-03 | **-10.37** |
| **GAMT** | 1.20E-02 | **-8.79** |
| **NDC80** | 1.93E-02 | **-8.50** |
| **ACSM3** | 1.56E-02 | **-7.12** |
| **NEIL3** | 3.59E-02 | **-6.75** |
| **CTXN1** | 5.70E-03 | **-6.54** |
| **SCAMP5** | 3.29E-03 | **-6.45** |
| **SDHAF4** | 2.14E-03 | **-6.30** |
| **HPDL** | 2.89E-02 | **-6.14** |
| **SPINT2** | 2.55E-02 | **-5.89** |
| **DNPH1** | 2.61E-02 | **-5.45** |
| **RASL10B** | 4.28E-03 | **-5.31** |
| **AURKB** | 6.85E-03 | **-5.20** |
| **INSM1** | 4.68E-02 | **-5.03** |
| **HILPDA** | 3.23E-02 | **-4.81** |
| **CENPV** | 7.59E-03 | **-4.73** |
| **COMTD1** | 3.89E-02 | **-4.47** |
| **APRT** | 2.02E-02 | **-3.97** |
| **SDC1** | 4.89E-02 | **-3.57** |
| **BHLHE40** | 1.70E-02 | **2.44** |
| **NEU1** | 6.04E-03 | **3.91** |
| **SDCBP** | 2.34E-03 | **4.09** |
| **STXBP1** | 2.41E-02 | **4.59** |
| **SNX30** | 7.90E-03 | **5.39** |
| **STX3** | 7.88E-04 | **7.78** |
| **CAMKK1** | 1.84E-02 | **9.19** |
| **SQSTM1** | 1.33E-02 | **12.03** |
| **EPHX1** | 4.82E-03 | **12.69** |
| **GADD45G** | 3.05E-02 | **12.89** |
| **RGS1** | 4.50E-02 | **15.93** |
| **UPP1** | 3.85E-02 | **17.67** |
| **LUCAT1** | 5.88E-06 | **21.88** |
| **LINC03016** | 1.04E-03 | **23.44** |
| **NQO1** | 3.21E-03 | **25.33** |
| **SERPINE1** | 2.47E-02 | **30.47** |
| **OSGIN1** | 2.90E-03 | **50.24** |

S2. B. IBR+S63845 Combination

| **Gene name** | **P-value (MINO-P,IBR_S63845 vs MINO-P,CON)** | **Fold change (MINO-P,IBR_S63845 vs MINO-P,CON)** |
| --- | --- | --- |
| **ABCG1** | 3.33E-04 | **-20.794** |
| **LCK** | 9.13E-03 | **-13.473** |
| **LINC02642** | 7.07E-03 | **-12.108** |
| **ADAMTS6** | 2.81E-03 | **-9.645** |
| **NIPSNAP1** | 3.00E-06 | **-9.554** |
| **PIF1** | 2.71E-02 | **-9.314** |
| **CCDC28B** | 6.38E-03 | **-8.513** |
| **TMEFF1** | 2.23E-02 | **-7.645** |
| **PVRIG** | 1.94E-03 | **-7.229** |
| **KLHL13** | 2.26E-08 | **-6.866** |
| **PAOX** | 2.12E-02 | **-6.235** |
| **LYAR** | 1.94E-03 | **-5.938** |
| **ICA1** | 8.68E-03 | **-5.306** |
| **DLGAP3** | 1.45E-04 | **-5.022** |
| **ATP7B** | 2.30E-02 | **-5.006** |
| **APOM** | 2.34E-02 | **-4.471** |
| **PACC1** | 4.33E-03 | **-4.223** |
| **WNK2** | 1.98E-03 | **-4.179** |
| **HMGA1P2** | 4.70E-02 | **-3.758** |
| **REEP2** | 4.53E-02 | **-3.614** |
| **MIPEP** | 8.07E-03 | **-3.564** |
| **ESD** | 2.83E-02 | **-3.477** |
| **CD4** | 3.28E-03 | **-3.434** |
| **ECE1-AS1** | 2.47E-06 | **-3.247** |
| **PEX7** | 4.98E-02 | **-2.829** |
| **EIF5AP4** | 3.26E-02 | **-2.714** |
| **RASA4DP** | 1.59E-03 | **-2.709** |
| **RDM1** | 1.09E-02 | **-2.444** |
| **OTX1** | 1.95E-02 | **-2.435** |
| **SLC2A4** | 3.22E-04 | **-2.435** |
| **ATP5F1C** | 1.69E-02 | **-2.331** |
| **PKD1L2** | 1.33E-02 | **-2.311** |
| **RHPN1-AS1** | 1.90E-03 | **-2.297** |
| **RDM1P5** | 9.94E-06 | **-2.061** |
| **PPP1R1B** | 2.99E-03 | **-2.061** |
| **VCAN** | 5.09E-04 | **-1.874** |
| **SNX5P1** | 8.51E-03 | **-1.812** |
| **LINC03062** | 2.41E-04 | **-1.587** |
| **ASIC1** | 3.28E-03 | **-1.437** |
| **RPL29P20** | 6.95E-05 | **-1.437** |
| **CD3E** | 3.60E-02 | **-1.437** |
| **MCIDAS** | 6.55E-04 | **-1.437** |
| **RAC1P4** | 3.44E-03 | **-1.374** |
| **CDKL4** | 1.25E-02 | **-1.187** |
| **MIR130AHG** | 5.23E-03 | **-1.187** |
| **GUCY2F** | 1.91E-02 | **-1.187** |
| **PRRT1B** | 5.52E-03 | **1.542** |
| **MSTO2P** | 9.32E-05 | **1.734** |
| **QPRT** | 1.90E-02 | **3.773** |
| **SIGLEC15** | 9.54E-03 | **5.154** |
| **CLEC2B** | 9.80E-05 | **7.138** |
| **FBXO32** | 8.14E-04 | **8.567** |
| **MT-ND2** | 4.70E-02 | **10.638** |
| **EPHX1** | 5.49E-03 | **12.078** |
| **GADD45G** | 2.98E-02 | **13.073** |
| **MT-CYB** | 2.05E-02 | **13.592** |
| **SQSTM1** | 8.71E-03 | **14.503** |
| **LINC01484** | 2.36E-03 | **15.003** |
| **MT-RNR1** | 1.75E-02 | **27.136** |
| **NQO1** | 1.88E-03 | **32.298** |
| **HMOX1** | 9.12E-03 | **48.832** |
| **SNORD3A** | 1.32E-02 | **156.844** |

S2.C. BTZ+YM155 Combination

| **Gene name** | **P-value (MINO-VR,BTZ_YM155 vs MINO-VR,CON)** | **Fold change (MINO-VR,BTZ_YM155 vs MINO-VR,CON)** |
| --- | --- | --- |
| **TERT** | 1.93E-05 | **-19.82** |
| **SPIB** | 1.99E-02 | **-18.38** |
| **FAM72A** | 4.82E-02 | **-15.98** |
| **HPDL** | 2.69E-03 | **-14.90** |
| **KCNA3** | 2.49E-02 | **-14.69** |
| **FAM72D** | 1.96E-02 | **-14.59** |
| **GAMT** | 4.64E-03 | **-12.56** |
| **INSM1** | 4.56E-03 | **-12.07** |
| **CD72** | 2.84E-02 | **-11.82** |
| **ACSM3** | 4.99E-03 | **-10.69** |
| **TMPO-AS1** | 1.83E-02 | **-9.08** |
| **PIMREG** | 1.74E-02 | **-8.55** |
| **NEIL3** | 2.66E-02 | **-7.68** |
| **NDC80** | 2.73E-02 | **-7.36** |
| **PXMP2** | 2.37E-02 | **-7.30** |
| **SDC1** | 5.95E-03 | **-6.72** |
| **CENPV** | 2.18E-03 | **-6.44** |
| **AURKB** | 3.77E-03 | **-6.07** |
| **SCAMP5** | 9.16E-03 | **-4.93** |
| **SDHAF4** | 5.91E-03 | **-4.91** |
| **ABCG1** | 3.72E-02 | **-4.45** |
| **CTXN1** | 2.46E-02 | **-4.28** |
| **HILPDA** | 4.70E-02 | **-4.22** |
| **CADM4** | 2.45E-02 | **-4.03** |
| **SDCBP** | 1.88E-02 | **2.75** |
| **BHLHE40** | 6.55E-03 | **2.85** |
| **NEU1** | 9.57E-03 | **3.55** |
| **STX3** | 5.29E-03 | **4.91** |
| **PSAP** | 4.01E-02 | **6.51** |
| **LUCAT1** | 4.60E-04 | **7.45** |
| **NQO1** | 2.36E-02 | **10.16** |
| **LINC03016** | 9.13E-03 | **10.21** |
| **SQSTM1** | 5.59E-03 | **17.61** |
| **UPP1** | 3.66E-02 | **18.27** |
| **GADD45G** | 4.01E-03 | **38.13** |
| **OSGIN1** | 3.08E-03 | **48.69** |

S2.D. BTZ+S63845 Combination

| **Gene name** | **P-value (MINO-VR,BTZ_S63845 vs MINO-VR,CON)** | **Fold change (MINO-VR,BTZ_S63845 vs MINO-VR,CON)** |
| --- | --- | --- |
| **IL10** | 3.38E-03 | **-29.246** |
| **CCDC28B** | 1.55E-03 | **-13.639** |
| **IGHD** | 8.10E-03 | **-12.100** |
| **LINC02642** | 1.45E-02 | **-9.111** |
| **ATP7B** | 7.11E-03 | **-7.269** |
| **ICA1** | 2.77E-03 | **-7.239** |
| **LYAR** | 1.49E-03 | **-6.322** |
| **ABCG1** | 2.49E-02 | **-5.093** |
| **ADAMTS6** | 4.66E-02 | **-3.949** |
| **PVRIG** | 3.09E-02 | **-3.500** |
| **CD4** | 3.30E-03 | **-3.430** |
| **NIPSNAP1** | 2.06E-03 | **-3.167** |
| **RSPH1** | 3.07E-03 | **-3.136** |
| **WNK2** | 9.17E-03 | **-3.132** |
| **RSPH14** | 4.87E-03 | **-3.061** |
| **PACC1** | 2.91E-02 | **-2.810** |
| **RASA4DP** | 1.30E-03 | **-2.780** |
| **SLC2A4** | 1.06E-03 | **-2.171** |
| **CFAP58** | 1.68E-02 | **-2.156** |
| **GUCY2F** | 2.22E-08 | **-2.031** |
| **KANK3** | 9.98E-03 | **-1.945** |
| **MST1P2** | 7.31E-04 | **-1.907** |
| **LINC02321** | 3.18E-02 | **-1.890** |
| **RDM1P5** | 5.94E-05 | **-1.843** |
| **MANSC1** | 6.14E-03 | **-1.776** |
| **TLK1P1** | 1.37E-02 | **-1.749** |
| **HIVEP2-DT** | 7.58E-04 | **-1.589** |
| **PFKFB1** | 3.76E-02 | **-1.576** |
| **KLHL13** | 2.42E-02 | **-1.562** |
| **VCAN** | 2.55E-02 | **-1.422** |
| **SINHCAFP1** | 1.36E-02 | **-1.375** |
| **MSTO2P** | 2.43E-02 | **-1.296** |
| **LINC03062** | 2.09E-02 | **-1.281** |
| **RPL29P20** | 2.04E-02 | **-1.187** |
| **CLEC2B** | 4.34E-03 | **3.490** |
| **QPRT** | 8.74E-04 | **8.161** |
| **NQO1** | 2.60E-02 | **9.717** |
| **FBXO32** | 9.89E-05 | **14.973** |
| **SQSTM1** | 3.92E-03 | **20.551** |
| **GADD45G** | 4.46E-03 | **36.037** |
| **HMOX1** | 4.32E-03 | **78.835** |
